# Amygdala subregional functional connectivity in Treatment-Resistant Depression

**DOI:** 10.1101/2024.09.22.24314177

**Authors:** Sheryl L. Foster, Ramon Landin-Romero, Sarah Lewis, Ana Rita Barreiros, Sophie Matis, Anthony Harris, Mayuresh S. Korgaonkar

**Affiliations:** Discipline of Medical Imaging Science, School of Health Sciences, Faculty of Medicine and Health, The University of Sydney, Sydney, NSW 2006, Australia; Department of Radiology, Westmead Hospital, Westmead, NSW 2145, Australia; School of Health Sciences, Western Sydney University, Penrith, NSW, 2751; Westmead Clinical School, Faculty of Medicine and Health, The University of Sydney, Sydney, NSW 2006, Australia; Brain Dynamics Centre, The Westmead Institute for Medical Research, Westmead, NSW, 2145, Australia; Brain and Mind Centre, The University of Sydney, Camperdown, NSW 2050, Australia; Specialty of Psychiatry, Sydney Medical School, The University of Sydney, Sydney, NSW 2006, Australia; The Black Dog Institute, Prince of Wales Hospital, Randwick, NSW 2031, Australia

**Keywords:** Treatment-Resistant Depression, amygdala, subregions, functional connectivity, high-resolution

## Abstract

**Background:** Treatment resistance impacts almost 50% of depression patients, with amygdala dysfunction being widely implicated. The amygdala is a complex amalgamation of subnuclei with diverse functions but fMRI studies have typically focussed on identifying whole rather than subregional amygdala functional connectivity. This study used high resolution 3T fMRI data to investigate subregional alterations that may differentiate treatment-resistant cohorts from healthy individuals and depressed patients who respond to treatment.

**Methods:** Resting-state fMRI data were obtained in 35 participants diagnosed with Treatment-Resistant Depression (TRD), 38 healthy control participants (HC), and 35 treatment-sensitive participants (TSD). Seed-based functional connectivity analyses of the three main subregions bilaterally (laterobasal, centromedial and superficial), as well as the whole amygdala, were performed and comparisons made between groups.

**Results:** We found **c**onnectivity differences in the right laterobasal amygdala subregion in TRD compared to both groups. TRD patients displayed hypoconnectivity to the right fusiform gyrus relative to HC whereas hyperconnectivity to the left inferior frontal gyrus relative to TSD was identified. No connectivity differences were found for the whole amygdala or any of the other subregions bilaterally.

**Limitations:** Modest sample size and cross-sectional study design are limitations. A causal relationship between functional connectivity alterations and treatment resistance cannot be established.

**Conclusion:** Altered connectivity of the right laterobasal subregion is a distinguishing feature of TRD. These alterations may underlie severe impairments in emotion processing and social functioning that are characteristic of TRD. These results emphasise the need for further investigation of the functional role of the amygdala subregions in depression.

## 1. Introduction

Of all patients diagnosed with Major Depressive Disorder, only around half will adequately respond to initial treatments based on antidepressant medications and targeted psychological interventions. These patients are considered Treatment-Sensitive (TSD) (Malhi et al., 2021; Scott et al., 2023). Approximately 30% of depression patients exhibit a lack of adequate response to initial pharmacotherapy plus at least one other pharmaceutical or alternative equivalent treatment and are diagnosed with treatment-resistant depression (TRD) (Ionescu et al., 2015; Voineskos et al., 2020). However, this figure could be as high as 55%, as it is well-recognised that the lack of a universal definition contributes to widely variable estimates of prevalence across treatment settings (McIntyre et al., 2023).

The neural mechanisms that distinguish TRD from TSD remain unclear and current diagnostic methods, predicated on a combination of patient self-reporting and monitoring of symptoms, are clearly suboptimal. Improved outcomes for TRD patients depend on further advancements in diagnosis rather than multiple trials of treatments which are ineffective for many patients (Malhi et al., 2019; McIntyre et al., 2023). Increasing access to clinical research Magnetic Resonance Imaging (MRI) systems has meant that advanced neuroimaging techniques such as functional MRI (fMRI) are now well-established in Major Depressive Disorder research (Kotoula et al., 2023). Limbic brain structures such as the amygdala have become a focus for mental health researchers. This is largely due to amygdala dysfunction being widely implicated in depression due to its role in emotion regulation and sensory processing (Drevets, 2003; LeDoux, 2000). Previous studies focusing on amygdala circuitry have clearly demonstrated the potential of fMRI to enhance our understanding of treatment responsiveness and remission in depressed cohorts (Goldstein-Piekarski et al., 2016; Williams et al., 2015).

This is supported by fMRI studies in treatment resistant depression that have reported altered activation and connectivity of the amygdala during both task and resting state fMRI (rs-fMRI) studies. However, findings across studies have been inconsistent, potentially due to diversity in acquisition and analysis techniques (Grehl et al., 2023; Kotoula et al., 2023). Incongruent fMRI task-based findings may be attributed to variations in fundamental task design across studies, whereas the rs-fMRI technique is easily replicated and should allow a more accurate comparison of findings across cohorts by analysing FC patterns during rest. However, reported findings of rs-fMRI FC patterns focusing on the amygdala in TRD have been inconsistent. Of the few FC studies comparing TRD with healthy controls (HC) there are reports of both hypoconnectivity (Ge et al., 2019; Lui et al., 2011; Wang et al., 2017) and hyperconnectivity (Siegel et al., 2021) of the amygdala with other brain regions. Similarly, in the only two studies found comparing FC in TRD and TSD, one reported amygdala hypoconnectivity in TRD relative to TSD (Lui et al., 2011) whilst the other found no significant differences between cohorts (Zhang et al., 2022).

Another important limitation in the field has been that the majority of studies comparing amygdala FC in TRD to HC and TSD cohorts have considered the amygdala as a single entity during data analyses, rather than a functionally diverse group of nuclei (Chen et al., 2020; Ge et al., 2019; Lui et al., 2011; Siegel et al., 2021; Vasavada et al., 2021; Wu et al., 2011). However, previous animal and human work has shown that the amygdala is not a simple homogenous structure; it is composed of nine functionally different groups of nuclei which are further grouped into three primary subregions, the laterobasal (LB), centromedial (CM), and superficial (SF) based on histological features and patterns of structural and functional connectivity (Figure 1) (Amunts et al., 2005; Bzdok et al., 2013; Kedo et al., 2018; Solano-Castiella et al., 2010). As well as being functionally diverse, the amygdala subregions also display asymmetric or lateralised connectivity (Ball et al., 2007; Bzdok et al., 2013; Kedo et al., 2018; Roy et al., 2009; Zhang et al., 2018). Acknowledging the need for greater accuracy in investigating and reporting subregional FC findings, parcellation techniques using high resolution 7 Tesla data have been developed (Eickhoff et al., 2005). The integration of probabilistic cytoarchitectonic maps into software analysis packages has enabled researchers to define separate standardised subregional regions-of-interest (ROIs)(Huang et al., 2024). This has facilitated more accurate reporting of localised FC alterations at a subregional rather than whole amygdala level.

**Figure 1.**
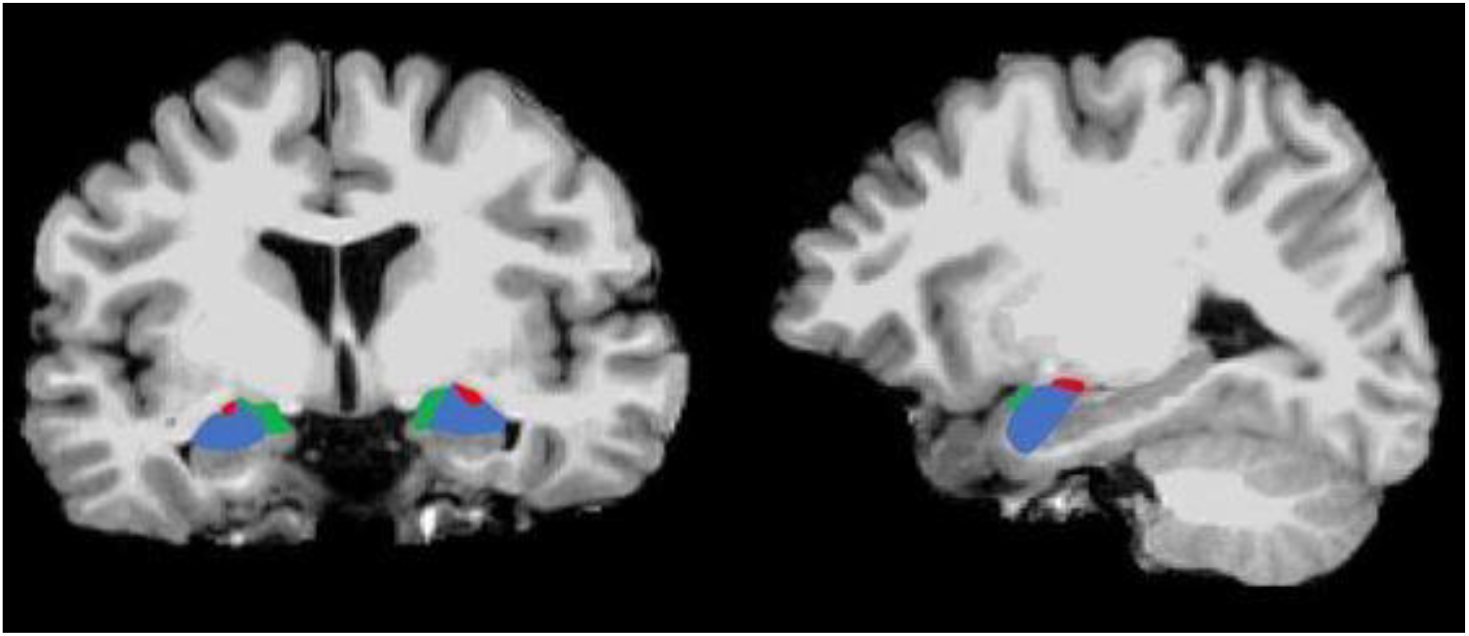
T1-weighted coronal (left), and sagittal (right) MR images depicting location of the amygdala and three main subregional ROIs. Superficial (SF = green), laterobasal (LB = blue) and centromedial (CM = red)

However, there is yet another confounder potentially contributing to the inconsistency of FC results. Spatial resolution of fMRI acquisition protocols, expressed as voxel volume in mm^3^, has remained largely unchanged, meaning the potential benefits of high-resolution fMRI data in imaging the subregions are predominantly unrealised (Foster et al., 2023; Olman & Yacoub, 2011). This is reflected in the handful of FC investigations of amygdala subregions in TRD to date that are primarily based on what is now considered standard (20-50mm^3^) and lower (> 50mm^3^) resolution data, likely contributing to the conflicting results being reported (Wang et al., 2017; Yuan et al., 2023; Zhang et al., 2022). Paradoxically, several TRD studies have acquired high-resolution data (< 20mm^3^) but reported FC alterations based on the whole amygdala rather than subregional ROIs (Nakamura et al., 2021; Vasavada et al., 2021) (Olman & Yacoub, 2011). While there has been one previous high-resolution subregional amygdala study in TRD vs HC that reported left SF hypoconnectivity (Batail et al., 2023), no previous study has evaluated subregional connectivity differences between TRD and TSD using high-resolution data. Given the limited understanding of the role of the amygdala subregions in TRD and the inconsistencies in results and imaging methods to date, there is clearly a need for further investigation.

The primary aim of this work was to examine alterations in resting-state FC of the amygdala in a TRD cohort compared with HC and TSD cohorts using high-resolution fMRI data and subregional amygdala ROIs. Based on previous studies, it was hypothesized that hypoconnectivity of the amygdala subregions would distinguish TRD from HC. We also hypothesized that amygdala subregional connectivity would differ between TRD and TSD. Further, as there are few reports of FC findings using both subregional ROIs and whole amygdala ROIs in the same cohorts, the secondary aim was to compare findings using both analysis methods.

## 2. Methods and Materials

### 2.1 Participants

Thirty-nine TRD patients, thirty-five TSD patients and thirty-eight healthy controls (HC) totalling 112 participants aged between 18 and 65 years were recruited for the study. Referencing the Structured Clinical Interview for the DSM-5 (SCID-5), all TRD and TSD participants met DSM-5 criteria for primary diagnosis of Major Depressive Disorder (APA, 2013). Symptom severity was characterized by a 17-item Hamilton Depression Rating Scale (HAMD-21) (Hamilton, 1960). TRD was defined as the presence of unremitting moderate to severe symptoms in patients who had undergone at least two trials of six weeks duration of adequate dosage of antidepressant medication of different pharmacological classes and a HAMD-21 score of 16 and above. TSD was defined as at least two weeks of symptom remission and a HAMD-21 score of nine and below. Healthy controls had no previous history of psychiatric illnesses as assessed using the SCID-5. Exclusion criteria included inability to provide consent, lack of proficiency in English language, current diagnosis of eating disorder or other psychiatric disorder, substance dependence for past three months, pregnancy, prior history of or current neurological disorder, prior brain injury, Electroconvulsive Therapy (ECT) or Transcranial Magnetic Stimulation in previous 6-month period, or contraindications for MRI. Further details relating to the cohorts can be found in a previous publication (Barreiros et al., 2022). Ethics approval for the research protocol was obtained from Western Sydney Local Health District Human Research Ethics Committee and written consent was obtained from all participants. Imaging studies were undertaken at Westmead Hospital Radiology Department, Sydney, Australia.

### 2.2 Data acquisition

All imaging data were acquired on a 3 Tesla Siemens Prisma MRI system in conjunction with VE11C software (Siemens Medical Solutions, Germany) and a 64-channel head/neck array RF coil for signal reception. Participants were instructed to remain still and awake whilst viewing a fixation cross projected onto a coil-mounted screen during 8 minutes of resting-state data acquisition. Imaging parameters for the functional 2D T2^*^-weighted GRE-EPI sequence were as follows: Repetition time (TR) = 1500ms; Echo time (TE) = 33ms; Field-of-view (FOV) = 255mm; Matrix = 104 × 104; Flip angle (FA) = 85°; Phase encoding direction = A to P; Total acceleration = 6(MB 3); 320 volumes ; 60 interleaved axial-oblique slices at 2.5mm thick (0mm gap) parallel to the AC-PC line were acquired covering the whole brain with an isotropic voxel size of 2.5mm^3^.

A 3D T1-weighted gradient echo sequence was also acquired to provide a high-resolution structural scan for use in the pre-processing workflow for normalisation. Imaging parameters were as follows: Repetition time (TR) = 2400ms; Echo time (TE) = 2.21ms; Field-of-view (FOV) = 256mm; Matrix = 288 × 288; Flip angle (FA) = 8°; Phase encoding direction = A to P; Acceleration (GRAPPA) = 2 ; 192 sagittal slices at 0.9mm thick parallel to the interhemispheric fissure were acquired covering the whole brain with an isotropic voxel size of 0.9mm^3^.

### 2.3 Imaging data analyses

Data analyses were performed using a combination of Matlab R2022b (The MathWorks Inc. Natick, Massachusetts), SPM12 (Wellcome Trust Centre for Neuroimaging, London, UK) and CONN functional connectivity toolbox v22b (http://www.nitrc.org/projects/conn/). Preprocessing of anatomical and functional images was performed using default settings in the CONN modular preprocessing pipeline (Nieto-Castanon, 2020b). In brief, the data underwent realignment, unwarping, coregistration and resampling for motion correction and magnetic susceptibility interactions. Outliers were excluded and a BOLD reference image was created for each subject. Data were then normalised into standard Montreal Neurological Institute space and segmented into grey matter, white matter and cerebrospinal fluid, then resampled to isotropic 2mm voxels. Functional data were smoothed with a Gaussian kernel of 6mm full width half maximum, denoised using a standard pipeline including regression of potential confounders (Nieto-Castanon, 2020a), followed by bandpass filtering (0.01-0.1 Hz). Data from four TRD participants were identified as having unacceptable levels of head motion and excluded from further analysis, leaving a total of 108 datasets.

First-level analyses were based on a priori selection of ROIs. Six amygdala subregional masks or ROIs were extracted from The JuBrain Anatomy Toolbox (Eickhoff et al., 2005). These ROIs, left and right centromedial (CM), laterobasal (LB) and superficial (SF) (Figure 1) were created using cytoarchitectonically-defined probabilistic maps from the JuBrain Cytoarchitectonic Atlas (Amunts et al., 2005).

A further two whole amygdala ROIs constructed from a combination of the three subregional masks (left and right) were also extracted for the comparison analysis. These were used as seed regions in CONN to explore whole-brain voxel-wise FC and to determine FC strength represented by Fisher’s z transform of the correlation coefficients from a weighted General Linear Model. This was computed by averaging the time course of all voxels in each seed ROI and correlating with the time course of every other voxel in the brain. Correlation coefficients were used in the group level analyses, performed using a General Linear Model. TRD and HC groups were compared using two-sample t-tests, followed by comparison of TRD and TSD groups. Statistical parametric maps were generated and results were thresholded using a combination of cluster-forming p < 0.001 voxel-level threshold and a familywise corrected p-FDR < 0.05 cluster-size threshold (Chumbley et al., 2010). Additionally, mean beta FC estimates were extracted from significant clusters identified in the primary analysis to further explore post-hoc secondary analyses.

### 2.4 Analyses of demographic and clinical factors

Comparison of all three groups was performed for age (one-way ANOVA) and gender (chi square test). Patient groups were also compared for clinical measures including depression severity (HAMD-21), quality of life (SOFAS), age at first depressive episode, length of time on antidepressant medications (independent sample t-tests) and history of ECT and suicidality (chi square tests). To further explore variances in neural measures between TRD and TSD and to confirm that these were not driven by clinical differences between the patient groups, a univariate GLM ANOVA was used to explore the effect of group (fixed factor) controlling for co-variates of age at first depressive episode, length of time on antidepressant medications, quality of life and HAMD-21 scores.

For the TRD group, the same clinical variables were also tested for potential correlations with FC values. Independent samples t-tests were undertaken to compare FC variances between those patients with and without a history of suicidal ideation, suicide attempts and ECT treatment.

All statistical analyses were undertaken using SPSS software version 29 (IBM-SPSS, 2012). All statistical tests were corrected for multiple comparisons and effects considered significant at p < 0.05.

## 3. Results

### 3.1 Demographic and clinical characteristics

Demographic and clinical characteristics data relating to the cohort are shown in Table 1. All groups were comparable for age and gender; as expected, the depressive profile of the TRD group was significantly worse in comparison with the TSD group. The TRD group exhibited greater severity of depressive symptoms, poorer functioning and a higher number of suicide attempts. Results for both group analyses are summarised in Table 2.

**Table 1.** Demographic and clinical characteristics of all study participants

|  | TRD (35) | TSD (35) | HC (38) | F/t/X<sup>2</sup> | sig |
| --- | --- | --- | --- | --- | --- |
| *Demographics* |  |  |  |  |  |
| Age, Mean ± SD [Min-Max] | 42.3 ± 14.1 [18.1–64.3] | 37.2 ± 11.0 [20.0–57.4] | 47.1 ± 14.3 [18.9–66.0] | n.s. | n.s. |
| Gender (M), N (%) | 14 (40) | 17 (48.6) | 17 (44.7) | n.s. | n.s. |
| *Clinical Profile* |  |  |  |  |  |
| Age of onset, Mean ± SD [Min-Max] | 26.97 ± 13.13 [8–53] | 21.66 ± 9.26 [8–50] | n.a. | n.s. | n.s. |
| HAMD-21 score, Mean ± SD [Min-Max] | 25.23 ± 6.46 [16–41] | 4.18 ± 3.10 [0–9] | n.a. | 15.819 | <0.001 |
| SOFAS score, Mean ± SD [Min-Max] | 73.91 ± 16.18 [40–100] | 89.58 ± 5.47 [78–95] | n.a. | –5.275 | <0.001 |
| History of ECT, N (%) | 10 (28.6) | 0 (0) | n.a. | 11.667 | 0.001 |
| History of Suicidal Ideation, N (%) | 28 (80) | 26 (74.3) | n.a. | n.s. | n.s. |
| History of Suicidal Attempt, N (%) | 19 (54.3) | 5 (14.3) | n.a. | 13.938 | <0.001 |
| Time on ADM (yrs), Mean ± SD [Min-Max] | 4.44 ± 3.71 [0.17–12] | 4.18 ± 4.16 [0.12–17] | n.a. | n.s. | n.s. |

Abbreviations: n.a. – not applicable, n.s. – not significant; SD – Standard Deviation; M – male; HAMD-21 – Hamilton Depression Rating Scale, 21 items; SOFAS – Social and Occupational Functioning Assessment Scale; ECT – Electroconvulsive Therapy; ADM – Antidepressant Medication; N – total number

**Table 2.**
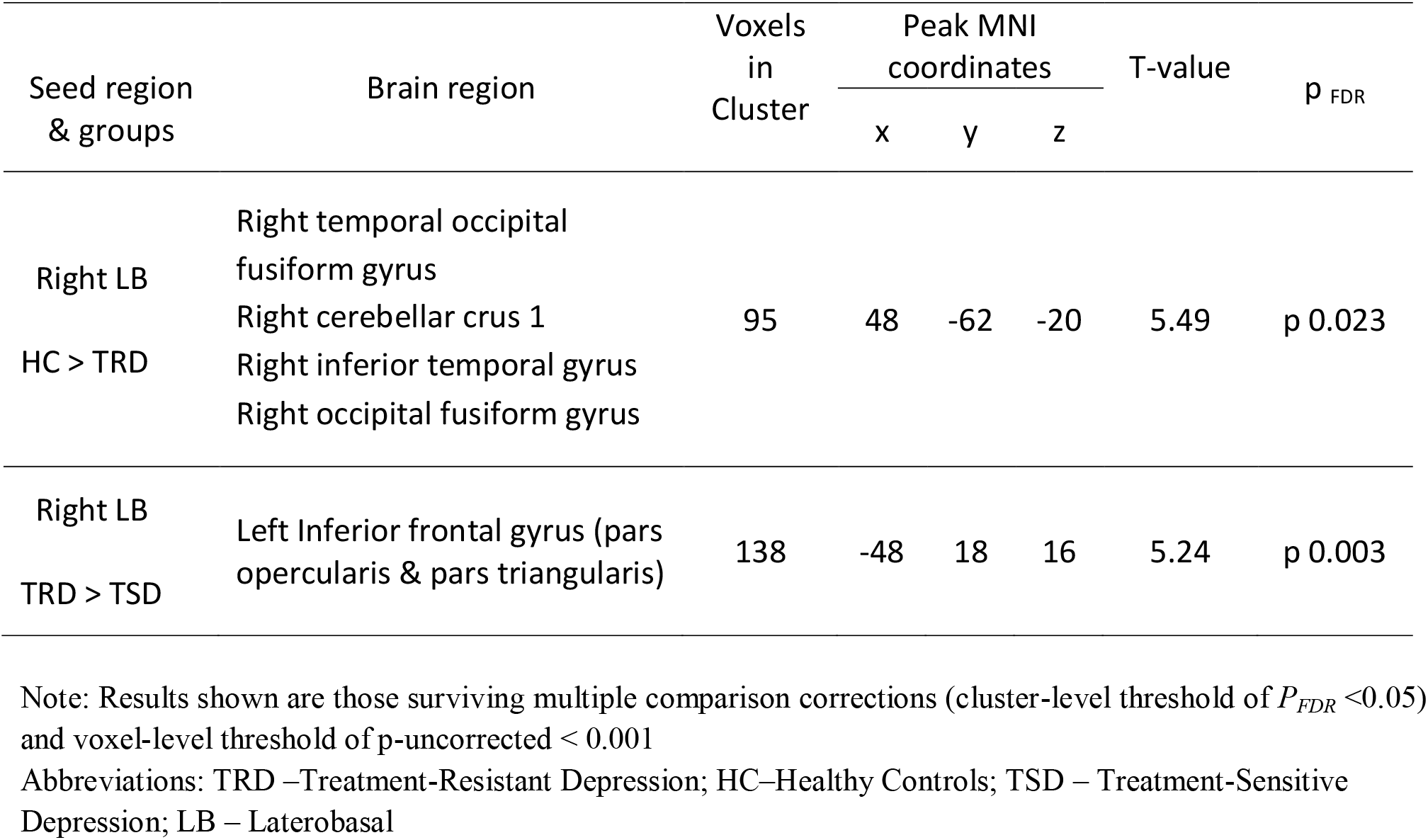
Summary of group results from functional connectivity analyses

### 3.2 Functional Connectivity Analyses -Healthy Controls vs TRD

Significant connectivity differences between patient groups were identified between the right LB and a right-sided cluster centred on the temporal occipital fusiform cortex with hypoconnectivity in the TRD group relative to HC (Figure 2). There were no FC group differences seen for any other subregion bilaterally.

**Figure 2.**
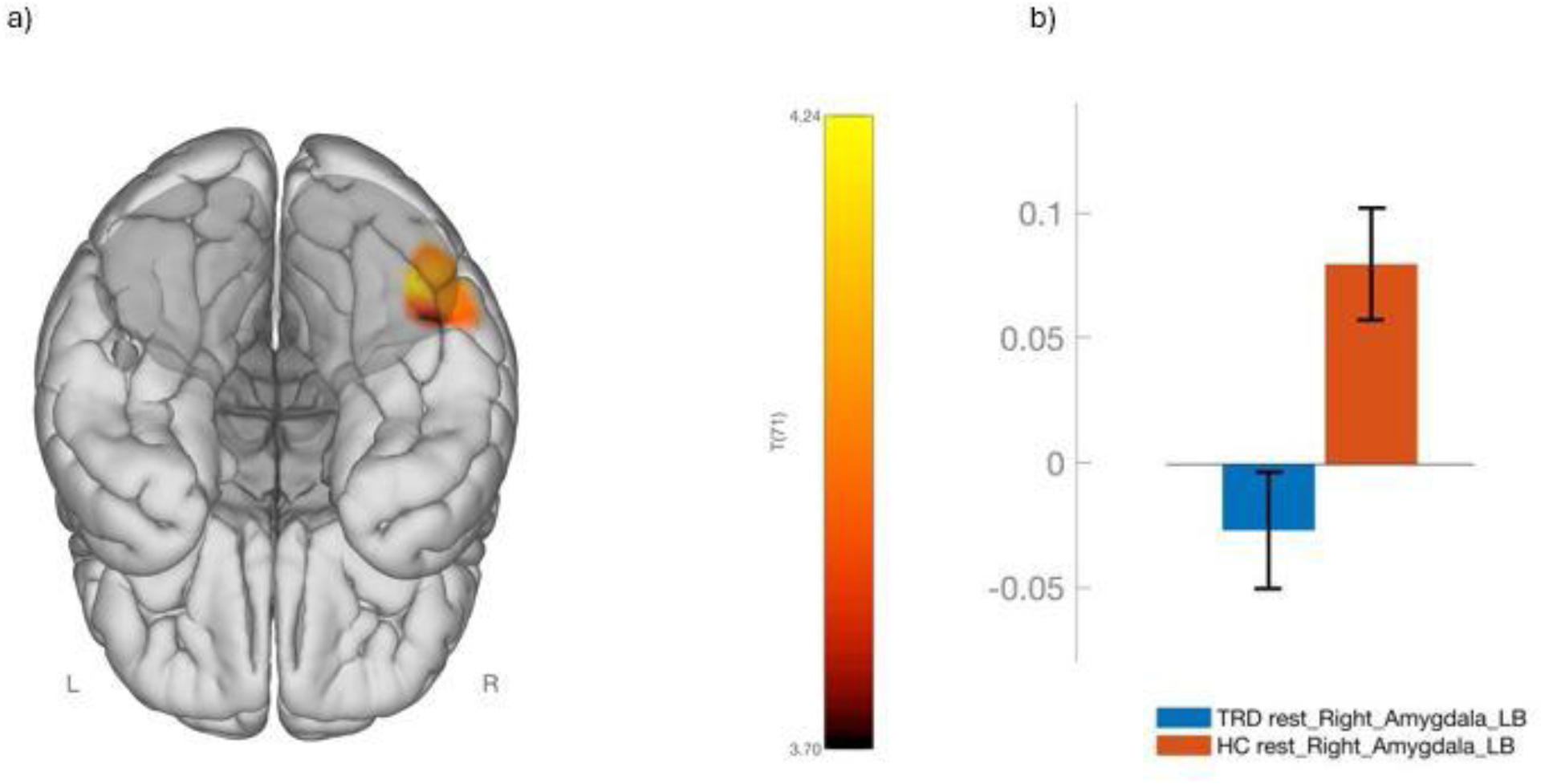
Functional connectivity (FC) differences between Treatment-Resistant Depression (TRD) and Healthy Control (HC) groups. a) significant cluster centred in the right temporal fusiform gyrus (inferior view) showed hyperconnectivity between right laterobasal (LB) amygdala subregion, with HC > TRD. b) mean FC of the right LB and right temporal fusiform gyrus for TRD and HC groups. The colour scale bar represents the strength of the t-statistic. The mean FC on b) represents beta values of FC between the two regions (95% CI).

No FC group differences were identified for the left or right whole amygdala ROI. However, at a less conservative threshold (uncorrected initial voxel threshold of p<0.005 with a p cluster corrected FDR threshold of 0.05) two clusters including parts of the hippocampus, cingulate gyrus, precuneus cortex and vermis, showed decreased FC with the right amygdala in the TRD group relative to HC.

### 3.3 Functional Connectivity Analyses – TRD vs TSD

Significant connectivity group differences were identified between the right LB and the left inferior frontal gyrus with hyperconnectivity in TRD relative to TSD (Figure 3). As observed in the previous analysis, there were no FC group differences seen for any of the other subregions bilaterally or for the left or right whole amygdala ROI, even at the less conservative threshold (uncorrected initial voxel threshold of p<0.005 with a p cluster corrected FDR threshold of 0.05).

**Figure 3.**
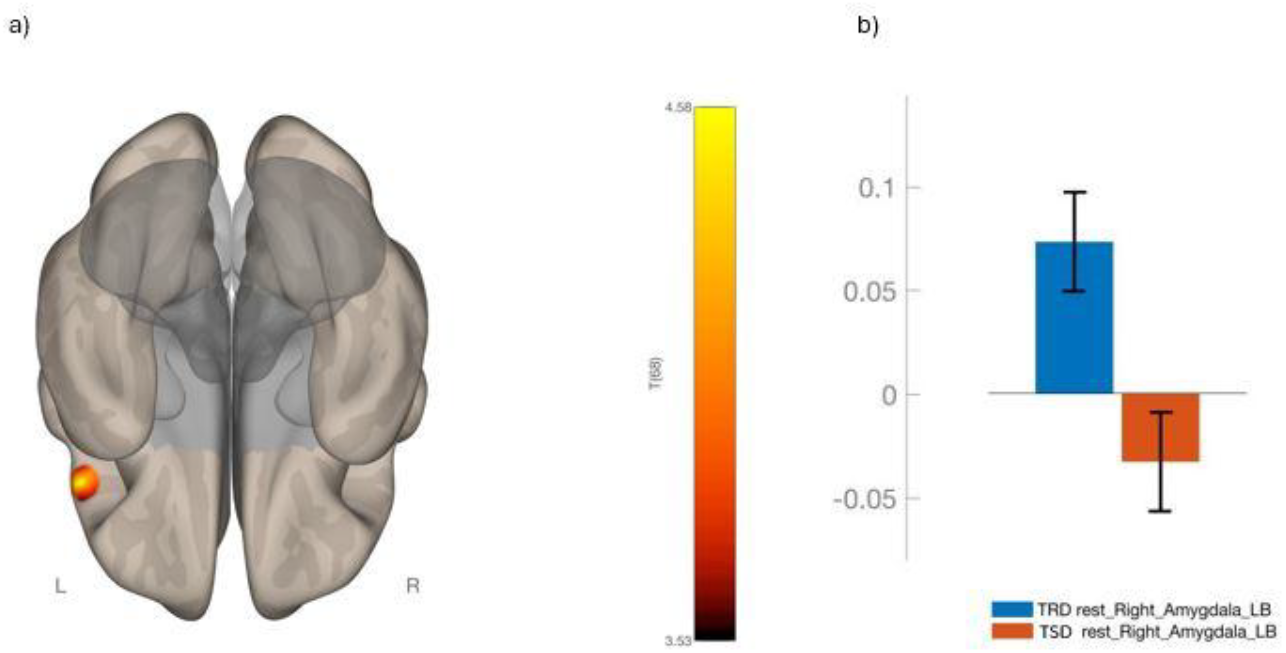
Functional connectivity (FC) differences between Treatment-Resistant Depression (TRD) and Treatment-Sensitive Depression (TSD) groups. a) significant cluster in the left inferior frontal gyrus (inferior view) showed hypoconnectivity between right laterobasal (LB) amygdala subregion, with TRD > TSD. b) mean FC of the right LB and left inferior frontal gyrus for TRD and TSD groups. The colour scale bar represents the strength of the t-statistic. The mean FC on b) represents beta values of FC between the two regions (95% CI)

### 3.4 Correlations between FC and clinical and demographic measures

Post hoc analyses were conducted by running a univariate GLM ANOVA to evaluate the potential of age at first episode, length of time on antidepressant medications, HAMD-21 and SOFAS scores to be contributing factors to the group differences in FC measures identified between TRD and TSD. After controlling for clinical measures, there was still a significant effect of group (F(1,44)=4.58, p=0.038), that is, there were no significant effects of these clinical measures on the FC differences identified between the right LB and the cluster in the left inferior frontal gyrus. In the TRD cohort, no significant effects were identified in testing for correlations with FC values using three clinical variables; patients with and without a history of suicidal ideation, suicide attempts and ECT.

## 4. Discussion

This study utilised high-resolution fMRI data to examine patterns of subregional amygdala functional connectivity in TRD relative to both depressed patients who respond to treatments (TSD) and healthy individuals (HC). As hypothesised, altered connectivity was identified in both group comparisons. Firstly, a pattern of hypoconnectivity was identified between the right amygdala LB subregion and the right fusiform gyrus in TRD relative to HC. Secondly, hyperconnectivity was found between the right amygdala LB and the left inferior frontal gyrus in the TRD cohort relative to TSD. These findings suggest that altered FC of the right LB subregion is a distinguishing feature of TRD.

These findings contribute to the limited work done so far in understanding the role of the amygdala and its subregions in TRD. This study highlighted that FC alterations of the right LB subregion are likely implicated in TRD. In comparison to HC, FC of the right LB was significantly decreased in TRD whereas it was significantly increased relative to TSD. To our knowledge, there are nine previous studies reporting amygdala resting state FC in TRD compared to HC, and only two comparing TRD and TSD. Based on a search of the literature, this study is the first to use both high resolution data and subregional ROIs to more accurately locate and report FC alterations in the amygdala subregions in a TRD cohort compared to TSD and only the second to compare TRD and HC cohorts.

The findings of the present study did not replicate previous findings in TRD relative to HC. Three of the six studies using whole amygdala ROIs reported hypoconnectivity of the right amygdala but to different brain regions (Chen et al., 2020; Ge et al., 2019; Vasavada et al., 2021). Another reported amygdala hyperconnectivity to the Default Mode Network (Siegel et al., 2021), and two found no significant amygdala FC differences (Lui et al., 2011; Wu et al., 2011). For the whole amygdala, we found differences between TRD and HC only at a less conservative threshold, with a pattern of hypoconnectivity for TRD to regions of the Default Mode Network, hippocampus and cerebellum. Of the three studies utilising subregional ROIs, two found left-sided hypoconnectivity of the SF, one to the left fusiform face area (Wang et al., 2017) and another to the left prefrontal cortex (Batail et al., 2023). The third study reported both hypoconnectivity of the right SF as well as hyperconnectivity of the right CM in their TRD cohort relative to HC (Zhang et al., 2022). Our study did not identify any connectivity differences related to the CM and SF subregions.

Very little work has been done in directly comparing TRD and TSD cohorts. The two previous studies identified in the literature had conflicting results that were also inconsistent with those of the present study. Significantly decreased connectivity between the left amygdala and left anterior cingulate cortex in TRD relative to TSD was identified in a study using low resolution data and whole amygdala ROIs (Lui et al., 2011) whereas no FC differences were identified in a study using standard resolution data and both whole and subregional ROI methods (Zhang et al., 2022). Our study also found no significant FC differences using whole amygdala ROIs in TRD relative to TSD.

Interestingly, within this small group of nine previous TRD studies that have reported on amygdala FC, cohort heterogeneity was evident. Underscoring the confounding nature of this issue, significant FC differences have also been shown within a TRD cohort in which a comparison was made between Anxious Depression and Non-anxious Depression subgroups (Yuan et al., 2023). In essence, the lack of congruency in FC findings across all these TRD studies is a hallmark that may be attributed not only to differences in acquisition and analysis methods but may be a reflection of the heterogeneity across TRD cohorts, supporting the premise that patients diagnosed with TRD may have little in common other than their lack of treatment response and providing a potential explanation for the limited success of current treatment regimes (Malhi et al., 2019).

The connectivity findings in the present study are interesting given what we know so far about the functional role of the LB subregion in the context of psychiatric disorders. Previous work has shown that the LB is involved in fear conditioning, regulates visual and auditory sensory input (Bzdok et al., 2013; Roy et al., 2009; Shen et al., 2019; Yang & Wang, 2017). It also acts as a connectivity hub and, as proposed by Pessoa, has an overarching “What is it?” function, meaning that it acts a decision-maker in relation to incoming information, particularly in terms of stimulus value (Pessoa, 2010). This subregion has also been shown to play a role in anxiety, social stress and negative regulation of social behaviour (Felix-Ortiz & Tye, 2014; Yang & Wang, 2017).

The amygdala and fusiform gyrus are linked structurally by the inferior longitudinal fasciculus and a relationship between the two has been established in previous task-based fMRI studies, supporting the role of both structures in facial perception (Chan et al., 2009; Edmiston et al., 2024; Frank et al., 2019; Müller-Bardorff et al., 2018). In particular, the right fusiform gyrus has been shown to be the primary contributor to visual processing including facial recognition and face perception (Ocklenburg & Güntürkün, 2018; Rangarajan et al., 2014; Zhang et al., 2016). Bidirectional communication between the right amygdala and right fusiform gyrus has been demonstrated, increasing in line with facial discrimination task load, in support of Pessoa’s “What is it?” hypothesis (Herrington et al., 2011; Pessoa, 2010). Interplay between the amygdala and the right fusiform gyrus has also been demonstrated in studies investigating depression using task-based fMRI (Chan et al., 2009; Sacu et al., 2023). Our findings are in line with a previous study that also reported decreased amygdala-fusiform FC in TRD relative to HC at baseline that subsequently increased after ECT treatment, although their findings were left-sided rather than right-sided (Wang et al., 2017). Our results of right LB amygdala hypoconnectivity with visual processing regions in TRD relative to HC parallel those in task-based studies that are suggestive of impaired bottom-up emotion processing of visual stimuli in the right amygdala (Ochsner et al., 2009; Ramasubbu et al., 2014). Given the known relevance of impaired facial processing in depressive symptoms, these findings may be indicative of a lesser known function of the amygdala-fusiform circuitry that could potentially be negatively impacting general social functioning in TRD cohorts (Akinci et al., 2022; Demenescu et al., 2010; Kupferberg & Hasler, 2023; Monferrer et al., 2023; Rutter et al., 2020).

The second key finding in TRD was hyperconnectivity of the right LB with the left inferior frontal gyrus compared to TSD. Part of the medial prefrontal cortex, the inferior frontal gyrus is implicated in several functions including semantic processing and social cognition. In particular, the left inferior frontal gyrus is thought to be responsive to the cognitive demands required during the execution of social tasks (Diveica et al., 2023) and also acts as a hub in the brain’s emotional circuitry (Li et al., 2021). Several previous studies using Amplitude of Low-Frequency Fluctuation and Regional Homogeneity analyses have demonstrated abnormal functional activity in the left inferior frontal gyrus in TRD relative to non-refractory Major Depressive Disorder, pinpointing this area as a potential differentiator between these cohorts (Guo et al., 2012; Sun et al., 2022; Wu et al., 2011). Our findings suggest that this could be due to connectivity related to the amygdala and, in particular, the LB subregion.

The top-down influence exerted on the amygdala by the medial prefrontal cortex is thought to be undermined by a range of chronic stressors causing structural amygdala neuronal remodelling and abnormal amygdala activation resulting in emotional disturbances (Liu et al., 2020; Zhang et al., 2021), with the LB subregion particularly implicated in relation to social stress and anxiety (Shen et al., 2019; Yang & Wang, 2017). There is growing evidence in human studies that the pathophysiology of depression and anxiety disorders may, in part, be related to stress-induced neuroinflammation promoting regional changes in brain function, structure and volume (Han & Ham, 2021; Milaneschi et al., 2021; Risbrough et al., 2022; Slavich & Irwin, 2014). Interestingly, around 25% of depression patients exhibit neuroinflammatory markers associated with treatment resistance and have poorer prognoses than those without inflammation (Hassamal, 2023).

Aside from cohort heterogeneity in TRD, there are also two potential technical explanations for lack of congruent FC findings across studies. There is considerable diversity in both data analyses and acquisition protocols, particularly in relation to the spatial resolution of fMRI data (Foster et al., 2023; Kotoula et al., 2023). The divergent findings in this study may reflect the use of high spatial resolution data acquired with voxel volumes of 15.6mm^3^ combined with subregional amygdala ROIs for data analyses. Of the eight previous TRD rs-fMRI studies identified, four reported whole amygdala findings using low resolution data (above 50mm^3^) whilst one used standard resolution data. Of the three studies reporting subregional findings, two used standard and only one used high-resolution data (Batail et al., 2023). Although larger voxels result in inherently higher signal-to-noise ratios, their use can be disadvantageous when imaging the amygdala. Due to the amygdala location deep in the temporal lobe near the sphenoid sinuses, intravoxel dephasing can occur during data acquisition, detrimentally affecting signal-to-noise ratio levels. Additionally, its position abutting the Basal Vein of Rosenthal, together with the diminutive size of the subregions, can result in confounding signals and partial volume effects which reduce signal accuracy and compromise the ability to resolve the very small subregional structures (Boubela et al., 2015; Olman & Yacoub, 2011). Further, the use of whole amygdala ROIs during data analyses may potentially confound findings. Zhang and colleagues reported both hypoconnectivity of the right SF and hyperconnectivity of the right CM in TRD compared with HC; however, their subregional results were not replicated in repeat analyses using a right whole amygdala ROI (Zhang et al., 2022). This was also the case in the present study, where the significant right LB subregional findings were unable to be replicated using the right whole amygdala ROI. These results support the premise that the common practice of considering the amygdala as a single structure may produce erroneous results, in part due to the potential for averaging of positive and negative subregional signals which are unable to be accurately interpreted using whole amygdala ROIs (Roy et al., 2009; Zhang et al., 2018).

## 5. Limitations and Future Work

There are several limitations to this study which may be overcome in future work. A causal relationship between treatment resistance and FC alterations cannot be implied due to the cross-sectional nature of the study. Severity of depression has the potential to be a confounding factor and, although the findings in this study held when controlling for depression severity in the TRD vs TSD comparison, resolving both these limitations in future work would necessitate a longitudinal approach in which depression patients are followed before and after TRD diagnosis. Additionally, as disparity in FC results is characteristic across the limited number of published TRD studies, replication of the current study results using fMRI data with similar or higher resolution, for example at 7T, would strengthen both the current findings and the case for acquisition of higher resolution fMRI data and a subregional amygdala ROI analysis approach in future studies. Although this study did not specifically evaluate anxiety as a coexisting condition across cohorts, this may have confounded the results as previous work has shown that altered connectivity of the subregions is implicated in anxiety (Etkin et al., 2009; Yuan et al., 2016). Future work could provide new insights into the role of anxiety as a potential contributor to symptom persistence in TRD.

## 6. Conclusion

This study showed that altered connectivity of the right LB subregion was a distinguishing feature of TRD. These results, obtained with high resolution data focusing on the amygdala subregions, emphasise the potential for improved techniques to advance our knowledge of the pathophysiological processes that underpin TRD. The study also provides a window into the highly specialised role of the right LB subregion and its complex interplay between different brain regions bilaterally. Review of the literature for this work revealed that heterogeneity -of cohorts, of data acquisition and of analysis techniques – is likely constraining our efforts to better understand the underlying neural mechanisms of TRD and future work may benefit from addressing these considerations.

## Data Availability

All data produced in the present study are available upon reasonable request to the authors

## CRediT authorship contribution statement

**Sheryl Foster:** Writing – review & editing, Writing – original draft, Visualization, Resources, Methodology, Investigation, Formal analysis, Data curation. **Ramon Landin-Romero:** Writing – review & editing, Methodology, Formal analysis, Supervision. **Sarah Lewis:** Writing – review & editing, Conceptualization, Supervision. **Ana Rita Barreiros:** Writing – review & editing, Resources, Methodology. **Sophie Matis:** Methodology, Writing – review & editing. **Anthony Harris:** Writing – review & editing, Resources, Project administration, Data curation, Conceptualization. **Mayuresh S. Korgaonkar:** Writing – review & editing, Supervision, Project administration, Funding acquisition, Conceptualization.

## Acknowledgement

This work was supported by The Westmead Charitable Trust Career Development Grant (to Sheryl Foster) and Takeda Pharmaceutical Company Limited [COCKPI-T](https://www.sciencedirect.com/science/article/pii/S2213158224000950?via%3Dihub#gp015) (Co-Create Knowledge for Pharma Innovation with Takeda) Research Grant [to Mayuresh Korgaonkar]), the National Health and Medical Research Council (NHMRC) (Grant No. [APP1087560](https://www.sciencedirect.com/science/article/pii/S2213158224000950?via%3Dihub#gp005) [to Mayuresh Korgaonkar]). None of the Takeda members had any specific role in the design and execution of the clinical study itself, but supported the conceptualization, data analysis and its interpretation.

## Data availability

The data that support the findings of this study are available from the corresponding author upon reasonable request.

